# Testing for tracing or testing just for treating? A comparative analysis of strategies to face COVID-19 pandemic

**DOI:** 10.1101/2020.06.01.20119123

**Authors:** Ricardo Knudsen

**Affiliations:** Independent Consultant

## Abstract

There is some consensus in Europe and Asia about high testing rates being crucial to controlling COVID-19 pandemics. There are though misconceptions on what means an effective high testing rate. This paper demonstrates that the rate of tests per detected case (Tests/Case) is the critical variable, correlating negatively with the number of deaths. The higher the Tests/Case rate, the lower the death rate, as this predictor is causally related to contact tracing and isolation of the vectors of the disease. Doubling Tests/Case typically divides by about three the number of deaths. On the other hand, the per capita testing rate is a poor predictor for the performance of policies to fight the pandemics. The number of tests per 1,000 inhabitants (Tests/1,000) tends to correlate positively with the number of deaths. In some cases, high levels of Tests/1,000 just mean an epidemic that ran out of control, with an explosion of cases that demands high testing rates just to confirm the diagnosis of the seriously sick. This study also demonstrates that an early tracing strategy, with a high level of Tests/Case, reduces combined costs of testing and hospitalization dramatically. Therefore, the common claim that tracing strategies are unaffordable by poorer countries is incorrect. On the contrary, it is the most adequate, both from the economic and humanitarian points of view.

## 1 Introduction

Countries affected by the COVID-19 pandemic followed two main conceptual strategies to face it. The most employed is also the most traditional, basically the same for the 1918 influenza pandemic. This approach is based on social distancing for the population, isolation and treatment for the infected ones. I’ll name it “Distancing, Isolating and Treating” strategy or DIT, for short. In DIT just the individuals who present symptoms of the disease are tested for COVID-19, for the sake of diagnosis.

The other main strategy is the “Trace, Test and Treat” or 3T, as named by the government of South Korea (1), where the policy was largely applied. In the 3T approach, confirmed cases have their contacts traced and tested, positive ones are quarantined. In 3T, a considerable fraction of the ones tested are not infected or infect but asymptomatic, presymptomatic or with light symptoms.

Other approaches, in general, would be placed somewhere in between DIT and 3T. The goal of this study was to understand what predictors characterize strategies and how they correlate with death rates. Focusing on the right indicators and concepts should help governments to design better policies against the pandemic.

Countries following 3T characteristics were the most successful in breaking the epidemic’s exponential growth, having less than 50 deaths per million inhabitants. In South Korea, the death rate was as low as five deaths per million by the time this article was written (last week of May 2020).

Most countries in Europe and North America followed the DIT strategy and in general had hundreds of deaths per million people. For instance, Spain and the United Kingdom had about 600 deaths/million. The state of New York, in the USA, had over 1,200 deaths/million, which is about 200 times the rate in South Korea and 1,000 times the one in Thailand.

One of the characteristics of 3T is a high testing level per case, or Tests/Case for short, for contact tracing. For this reason, 3T was often characterized by experts and the media as a “mass testing strategy”. Although this is true in the right context, considerable misconceptions have been created around it.

Due to the idea of “mass testing”, countries’ policies started to be evaluated in terms of the number of tests performed by the number of inhabitants, for instance the number of tests per 1,000 people (or Test/1,000, for short). As this paper demonstrates, either there is no correlation between Tests/1,000 and Deaths/Million people or there is a positive correlation. In other words, countries with higher Deaths/Million rates tend to have higher Tests/1,000 levels. These are in general the ones that followed the DIT strategy.

Another misconception is that the 3T strategy is about testing all (or most of) the population of a country. Consequently, some claim that 3T is unfeasible and a waste of public resources. This is of course a mischaracterization of 3T, and this paper shows that it indeed can reduce testing efforts and save public funds. As a matter of fact, DIT tends to be the mass testing strategy, as the number of tests grows exponentially to follow the explosion of cases.

The principal analysis for this study was made on data for 40 northern countries and selected states of the USA. Paper also presents a glimpse of nine tropical or south hemisphere countries, which were hit later by the pandemic and during a warmer season.

In this paper a high level of Tests/Case will be taken as an indication of some degree of 3T strategy. Data shows that countries that were testing only for diagnosis needed typically between 3 to 6 Tests/Case. Therefore, the ones testing considerably over that level were assumed to be testing individuals with light or no symptoms, trying to identify and isolate most of the disease’s vectors. Of course, not all those countries employed the same sophisticated technology and organization as in South Korea, nor reached the desired level of testing. But as this paper demonstrates, any increase in Tests/Case indicator pays off in terms of deaths reduction.

## 2 Methods

For the leading group analyzed, northern countries, I selected a set with similar characteristics to minimize the influence of effects other than their testing and tracing strategies. Criteria of choice for the northern countries were:

- Countries with high extension or population were excluded, as non-homogeneity would bring too much complexity into the analysis. Populations of selected countries lie between to 90 million people. Examples of countries excluded are the USA, China, Russia and Japan.
- To consider more information about North America, the northern states of the United States were included in the analysis, as equivalent to countries. American states have some administrative autonomy and indeed followed different practices towards the epidemic. Just the states with populations over 5 million people were considered. For simplicity, in this paper they will be referred to only as “countries”.
- All countries are above latitude 36 degrees north, which roughly corresponds to the Strait of Gibraltar. Therefore, all Europe, northern USA states and South Korea are in this group. That assures the epidemic started in winter or early spring, avoiding seasonal effects.
- All the countries and US states in the study adopted measures of social distancing or lockdowns for a specified period.
- Belgium was not included, as they count deaths for all those with typical symptoms, not only those confirmed by testing (2). This might have inflated deaths as compared to other countries.
- The analysis was made with the number of deaths rather than the number of cases, as it was more reliable. As demonstrated by Ricciardi, Verme, and Serajuddin (3), the apparent number of cases detected is a function of the testing rate. DIT highly underestimates the number of cases, as only those with symptoms are detected, while the number of deaths tends to be closer to reality. All the countries and US states were compared in the 6^th^ week after reaching about five deaths. Counting starts in the day the number of deaths is the closest to five (for instance, it could be 4 or 6). Just countries which had at least six weeks by May 9^th^ were considered.

On the last day of the 6^th^ week, indicators Tests/Case, Tests/1,000 and Deaths/Million were calculated from primary data. For Europe and South Korea, information is from the websites Worldometer (Total Deaths, Total Cases and Population) (4) and Our World in Data (Total Tests) (5). For the USA states, all the data is from the website “The COVID Tracking Project” (6).

Data for the tropical and southern hemisphere countries were obtained directly from the “Our World in Data” website. Choice’s criteria were as for northern countries, with two differences:

- South hemisphere is more economically heterogeneous than the northern countries chosen, which are mostly medium to high-income ones. Therefore, to avoid substantial economic effects in the comparisons, I limited the GDP per capita between 10,000 to 30,000 US dollars, in purchase power parity (7).
- The pandemic developed at a slower pace in the southern countries. To obtain more representative data for those countries, they were taken at eight weeks after five deaths, instead of six weeks.

Regression analysis was made with the statistical software Minitab 18.

## 3 Results

### 3.1 Northern Countries and US states

Tables 1 and 2 show data for the northern countries and the USA states used in the regression analysis. The column “Date 5 deaths” is the starting counting date and “Date 6 weeks” indicates when data was taken for regression analysis.

**Table 1.** Data for Countries

| Country | Date 5 deaths | Date 6 weeks | Tests/ Case | Testes/ 1,000 | Deaths/ million |
| --- | --- | --- | --- | --- | --- |
| Austria | 3/18 | 4/29 | 16.1 | 27.5 | 64 |
| Bulgaria | 3/27 | 5/8 | 29.0 | 7.8 | 12 |
| Canada | 3/16 | 4/27 | 14.8 | 19.0 | 72 |
| Czechia | 3/25 | 5/6 | 34.9 | 26.0 | 24 |
| Denmark | 3/18 | 4/29 | 20.0 | 31.1 | 76 |
| Finland | 3/25 | 5/6 | 19.4 | 19.5 | 44 |
| France | 3/4 | 4/15 | 3.6 | 7.4 | 263 |
| Germany | 3/12 | 4/23 | 15.3 | 28.0 | 67 |
| Greece | 3/17 | 4/28 | 27.2 | 6.7 | 13 |
| Hungary | 3/21 | 5/2 | 27.0 | 8.2 | 35 |
| Ireland | 3/22 | 5/3 | 9.2 | 39.9 | 264 |
| Italy | 2/24 | 4/6 | 5.4 | 11.9 | 273 |
| Netherlands | 3/11 | 4/22 | 5.5 | 11.1 | 237 |
| Norway | 3/18 | 4/29 | 22.0 | 31.2 | 38 |
| Poland | 3/17 | 4/28 | 25.4 | 8.2 | 16 |
| Portugal | 3/19 | 4/30 | 17.3 | 41.9 | 97 |
| Romania | 3/22 | 5/3 | 14.5 | 9.9 | 41 |
| Serbia | 3/25 | 5/6 | 12.0 | 13.4 | 23 |
| S. Corea | 2/22 | 4/4 | 44.8 | 8.9 | 3 |
| Spain | 3/5 | 4/16 | 5.2 | 20.6 | 409 |
| Sweden | 3/15 | 4/26 | 6.4 | 11.8 | 217 |
| Switzerland | 3/11 | 4/22 | 8.5 | 27.7 | 174 |
| Turkey | 3/19 | 4/30 | 8.6 | 12.2 | 38 |
| UK | 3/9 | 4/20 | 3.1 | 5.7 | 281 |
| Ukraine | 3/25 | 5/6 | 10.9 | 3.3 | 7 |

**Table 2.** Data for USA states

| Country | Date 5 deaths | Date 6 weeks | Tests/ Case | Testes/ 1,000 | Deaths/ million |
| --- | --- | --- | --- | --- | --- |
| Colorado | 3/22 | 5/3 | 4.9 | 13.9 | 144 |
| Illinois | 3/20 | 5/1 | 5.1 | 22.5 | 194 |
| Indiana | 3/21 | 5/2 | 5.4 | 15.5 | 183 |
| Maryland | 3/27 | 5/8 | 5.0 | 25.2 | 258 |
| Massachussets | 3/22 | 5/3 | 4.6 | 45.7 | 581 |
| Michigan | 3/21 | 5/2 | 4.7 | 20.2 | 402 |
| Minnesota | 3/28 | 5/9 | 9.8 | 18.8 | 99 |
| Missouri | 3/24 | 5/5 | 10.7 | 15.5 | 61 |
| New Jersey | 3/18 | 4/29 | 2.1 | 27.2 | 762 |
| New York | 3/15 | 4/26 | 2.8 | 41.0 | 872 |
| Ohio | 3/22 | 5/3 | 7.5 | 12.8 | 89 |
| Pennsylvania | 3/23 | 5/4 | 4.9 | 19.2 | 192 |
| Virginia | 3/23 | 5/4 | 5.8 | 13.1 | 80 |
| Washington | 3/2 | 4/13 | 12.6 | 17.5 | 69 |
| Wisconsin | 3/23 | 5/4 | 10.8 | 15.2 | 58 |

For Tests/Case versus Deaths/Million regression analysis, a power-law function was chosen, as it fitted better the results.

Figures 1A and 1B show regression results for Tests/Case vs. Deaths/Million, in a linear scale and in a double logarithmic scale, respectively. As we see, there is a strong negative correlation between the two variables, with Pearson’s correlation coefficient R= −0.859, and the determination coefficient R^2^ = 74%. It is remarkable that just one factor, Tests/Case, can explain 74% of the variance in Deaths/Million for all those countries.

Equation from regression analysis relating the two variables is: (Deaths/M) = 2326 x (Tests/Case)’’^45^

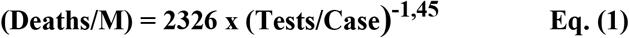

Equation (1) informs that doubling the number of tests per positive case, divides by 2.7 the number of deaths per million inhabitants, as an average.

**Figure 1A.**
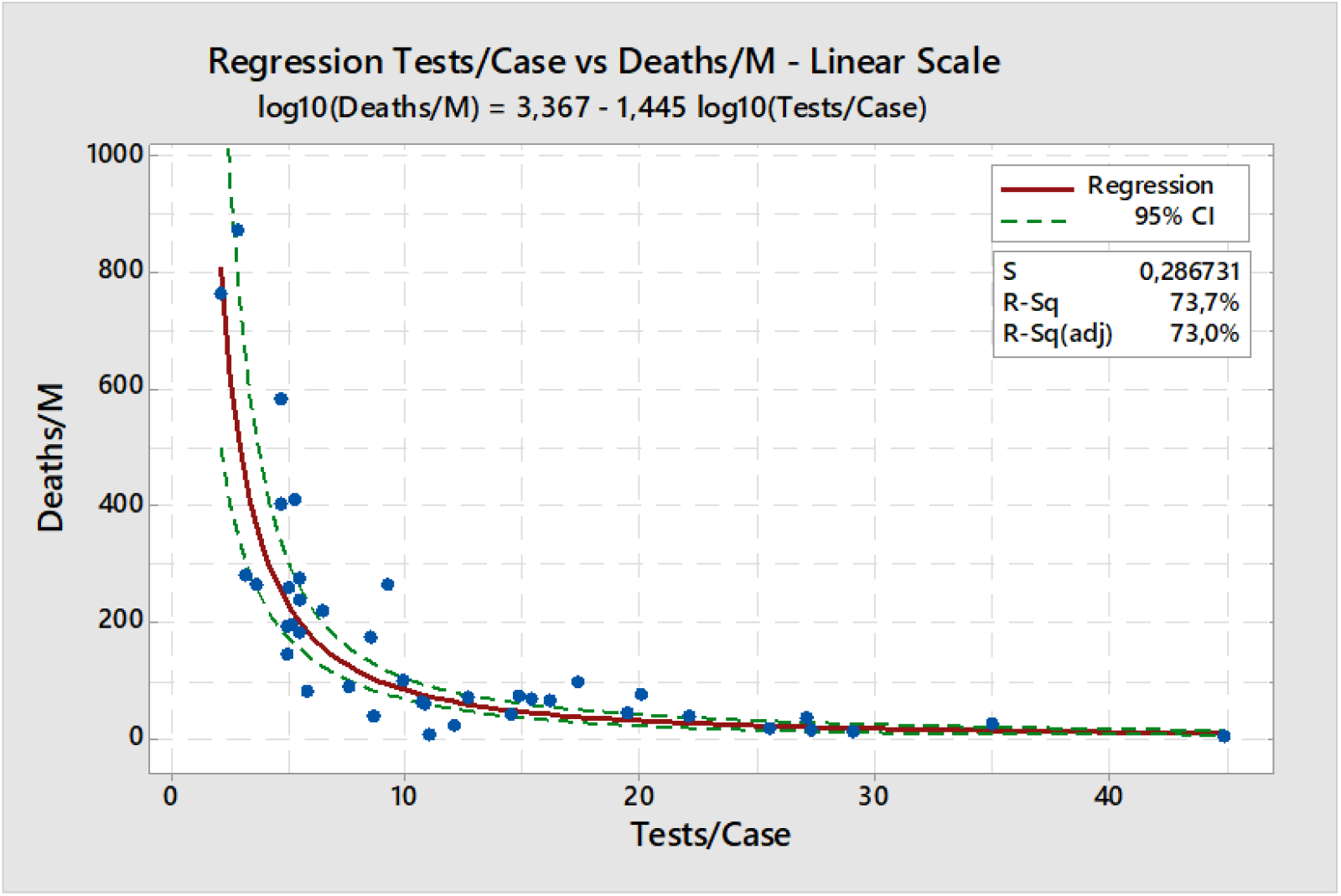
Regression Tests/Case vs. Deaths/Million - All Northen Countries - Linear scale.

**Figure 1B.**
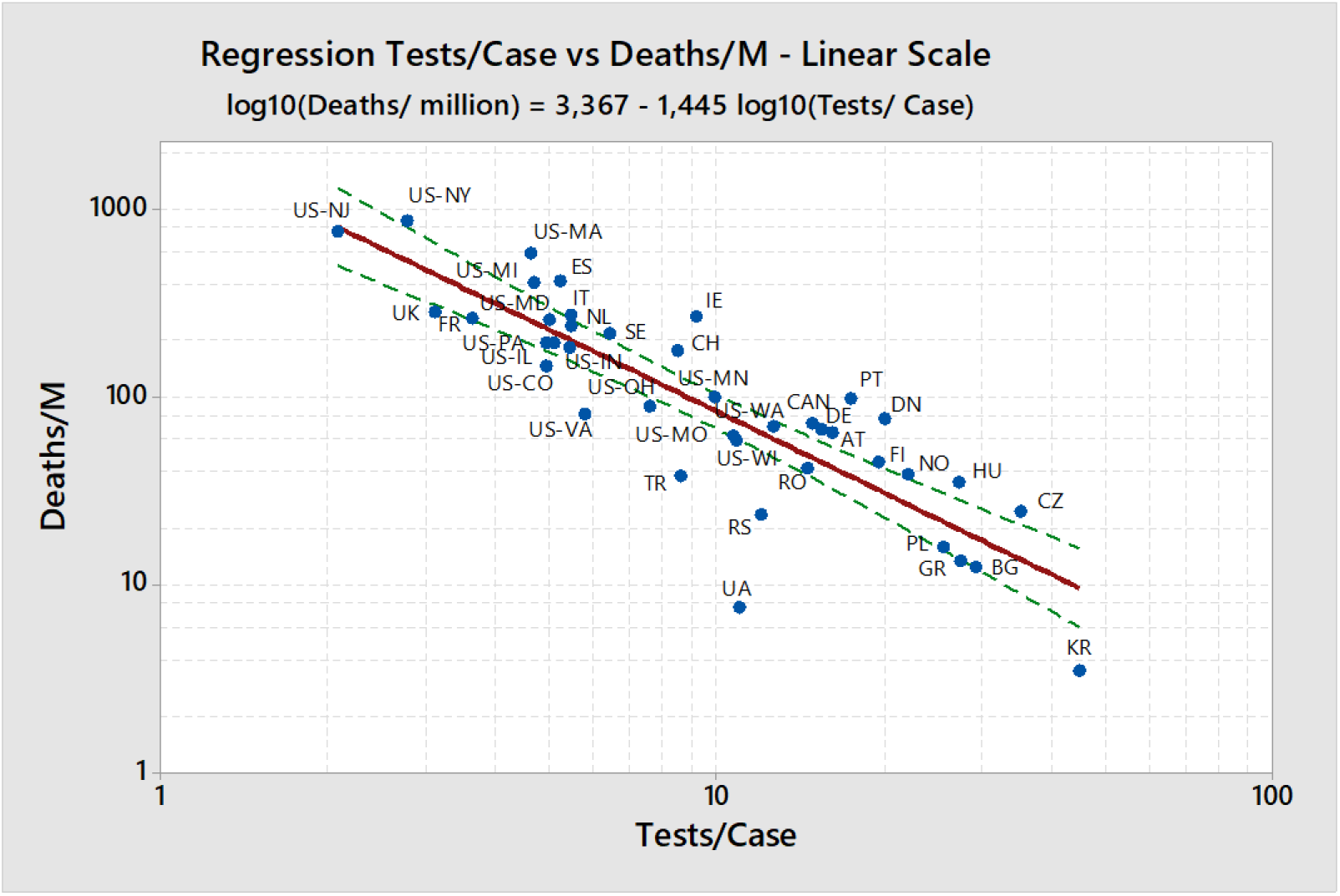
Regression Tests/Case vs. Deaths/Million - All Northen Countries - Log-Log scale.

Regression results for Tests/1,000 vs. Deaths/Million people are shown in Figure 2. As it is clear from data points dispersion, correlation is quite weak, and the determination coefficient is only 25%. Interesting to remark that the correlation, although weak, is positive, meaning the trend is that the higher the tests/1,000, the higher the death rate.

It is evident that Tests/1,000 is not a variable influencing death rate, being mostly a consequence of the number of cases, as it will be discussed later.

**Fig. 2.**
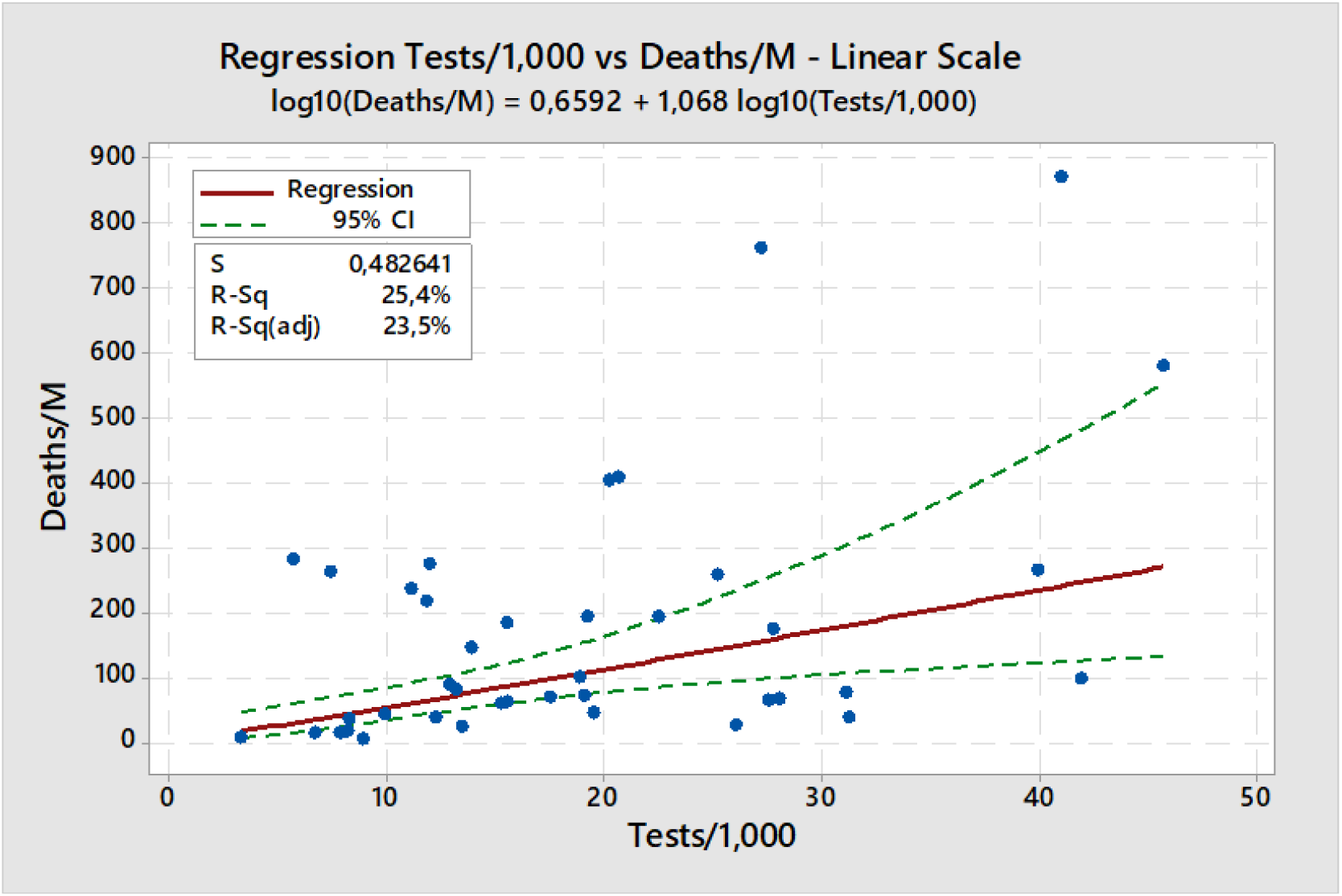
Regression Tests/1,000 vs. Deaths/Million - All Countries - Linear scale.

An alternative analysis was made, by reducing the group to Canada and the states of the USA. The idea was to analyze a more homogeneous group in terms of demographics, culture, climate, health characteristics, health services etc. For this regression, only data in Table 2 and from Canada were considered.

Regression for Tests/Case vs Deaths/Million is shown in figure 3. The equation representing the correlation is:

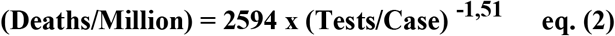

Correlation is negative, as expected, and indicates that doubling the number of tests per case divides the number of deaths per million by 2.84, in a fair agreement with the previous regression with all northern countries. Correlation, as expected, is higher, the determination coefficient is 79%.

**Figure 3.**
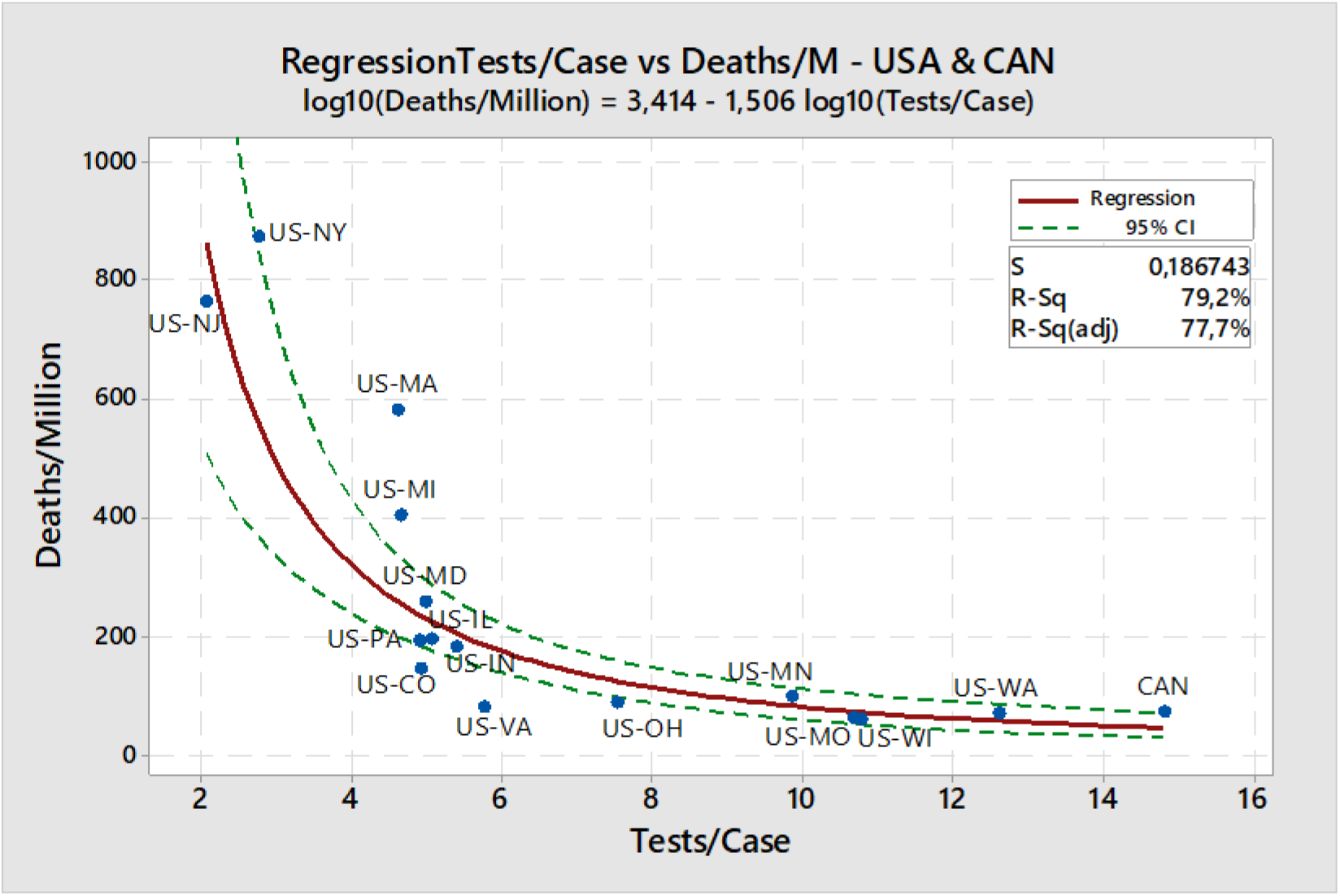
Regression Tests/Case vs. Deaths/Million - USA States & Canada - Linear scale.

In this case, regression for Tests/1,000 vs. Deaths/Million, shown in figure 4, is quite different. Contrarily to the case involving all countries, the correlation here is strong, with a determination coefficient of 69%. Once more it does not mean that higher Tests/1,000 is causing more deaths, but just that this is a consequence of the exponential growth of cases and deaths.

**Figure 4.**
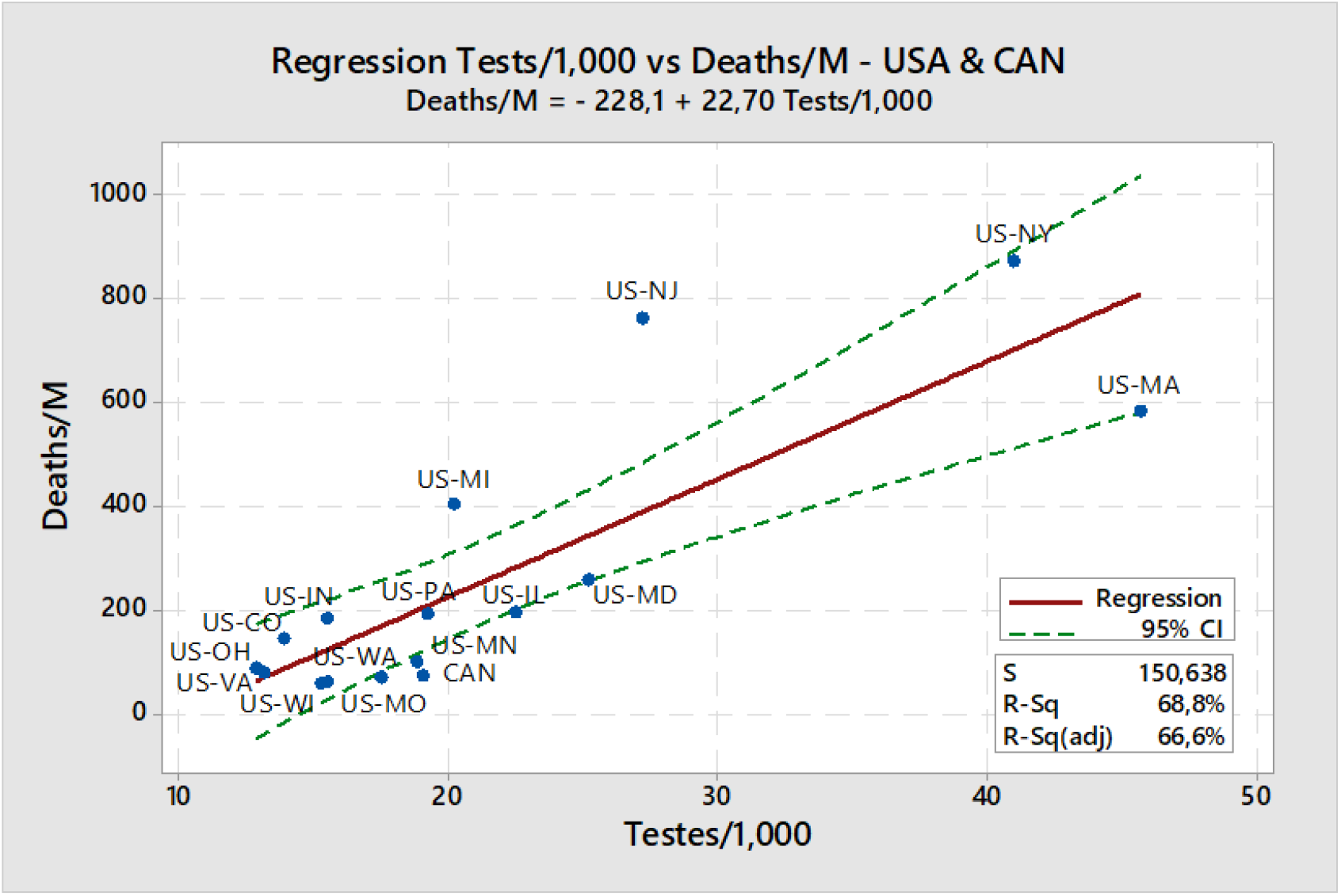
Regression Tests/1,000 vs Deaths/Million - USA States & Canada - Linear scale.

### 3.2 Southern Countries

By the time this work started, it was already clear that the pandemics in South America was following a different pattern, progressing more slowly. Possible reasons for that will be discussed later, but this is the main reason regression for southern and tropical countries was made separately.

Tunisia was included in the group, although not being in the tropical region or in the south hemisphere, as it fits the characteristics of medium-income rate, warmer climate and being hit latter by the pandemics. Tunisia is also an interesting case of a lower middle-income rate country which so far is being able to reduce death rates with a 3T-Like strategy.

As did previously, countries with larger populations and territories were not considered, like Brazil and India, for instance. But for Brazil, the state of São Paulo, which was the epicenter of the pandemics in the country, was included as a “country”. Having a population of 45 million people and a per capita GDP of about 26,000 dollars in ppp, São Paulo is quite comparable to the other countries selected.

Table 3 shows data for chosen southern countries, including data for GDP per capita in ppp.

**Table 3.** Data for southern countries

| country | GDP/<br>capita<br>(kUS\$) | Date 5<br>deaths | Date 6<br>weeks | Tests/<br>Case | Testes/<br>1,000 | Deaths/<br>million |
| --- | --- | --- | --- | --- | --- | --- |
| Argentina | 20.0 | 3/23 | 5/18 | 11.8 | 2.3 | 8.7 |
| Chile | 27.2 | 3/27 | 5/22 | 7.7 | 23.1 | 26.6 |
| Colombia | 16.3 | 3/26 | 5/21 | 12.6 | 4.4 | 12.1 |
| Ecuador | 11.9 | 3/19 | 5/14 | 1.5 | 2.5 | 160.9 |
| Peru | 15.4 | 3/21 | 5/16 | 1.1 | 2.9 | 88.4 |
| South Africa | 14.0 | 3/31 | 5/26 | 28.4 | 9.5 | 5.3 |
| Thailand* | 21.4 | 3/27 | 5/22 | 108.7 | 4.7 | 0.8 |
| Tunisia | 13.1 | 3/25 | 5/20 | 43.4 | 3.8 | 3.98 |
| São Paulo** | 25.7 | 3/19 | 5/14 | 3.3 | 13.7 | 102.2 |

\* Data from 5/15, last available.
\*\*The government of São Paulo (SP) does not inform regularly testing data. In a previous version of this article, Test/Case was 1.2 for SP, being the information available then (28). One month later, the government of SP informed that their data did not include tests from private laboratories, which would almost triple the rate. Therefore, I preferred to use this new data from June 15, 2020 (29), the only available with full information, which reflects better the testing rates in São Paulo.

Regression for Tests/Case vs. Deaths/Million is displayed in figure 5. For this group of countries, correlation is also negative but remarkably strong, with a determination coefficient of 95%. The regression equation is represented below:

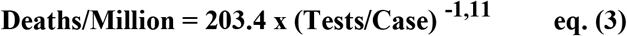

Equation 3 shows that there is a strong effect of testing per case rate on death rate. Interestingly though, this effect is slightly lower than for the northern countries. Doubling the number of tests per case would “only” about halve the number of deaths, compared to dividing by almost 3 for the northern countries.

**Figure 5.**
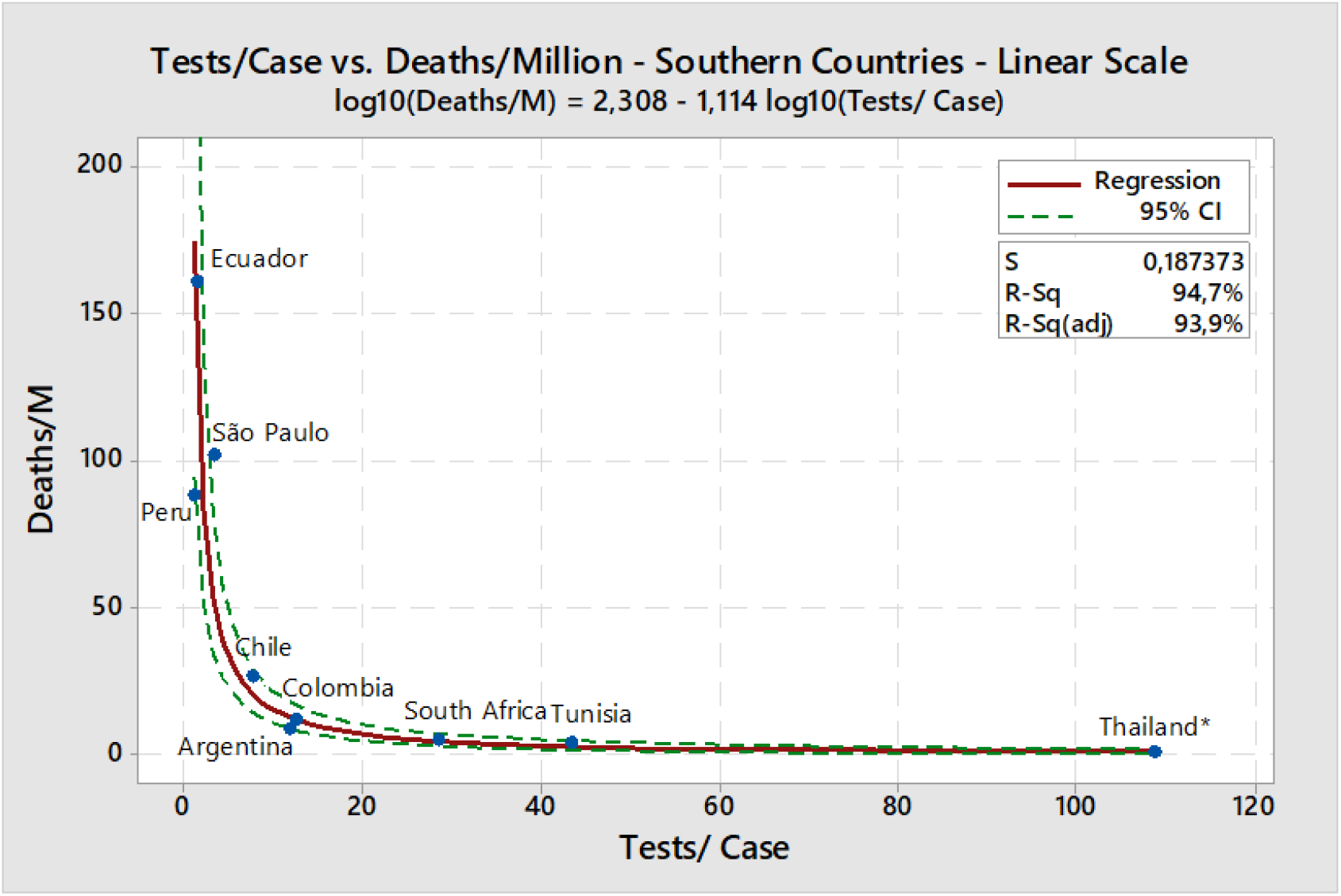
Regression Tests/Case vs. Deaths/Million - Southern Countries - Linear scale.

For the southern countries, the correlation between Tests/1,000 and Deaths/Million is even weaker than for the northern countries, in fact nonexistent. As it is shown in figure 6, datapoints are quite scattered, without any trend, and the determination coefficient is close to zero.

**Figure 6.**
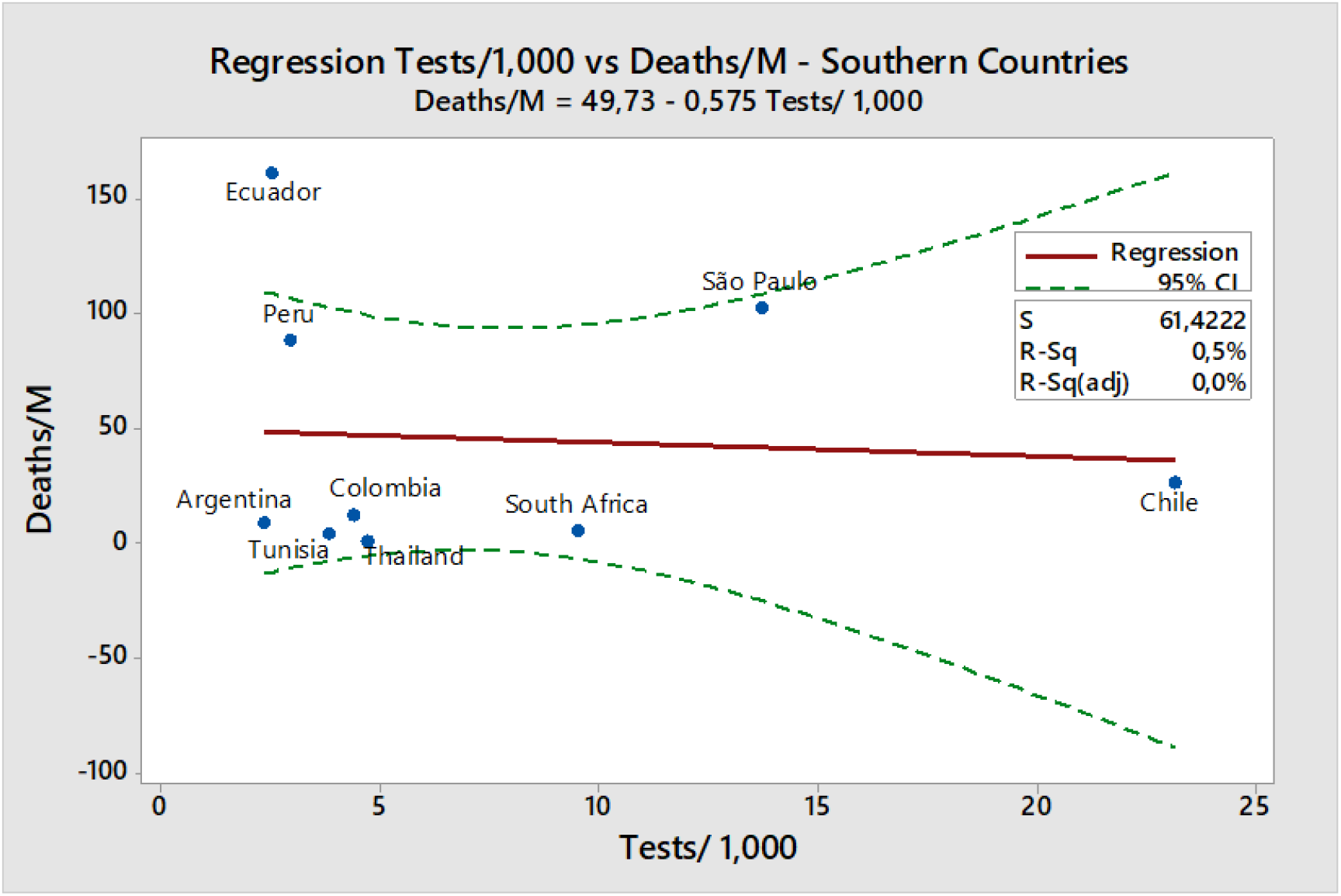
Regression Tests/1,000 vs. Deaths/Million - Southern Countries.

## 4 Discussion

### 4.1 General aspects

Regression results for both northern and southern countries show that the predictor for death rates related to testing is Tests/Case and not Tests/1,000 people. Therefore, Tests/Case is the best indicator to support establishing and evaluating policies against the pandemic.

To evaluate better the impact of Tests/Case and contact tracing in the pandemics, let’s make some simple simulations. Consider a country following a typical DIT strategy in the beginning of the epidemic. What would change if the health authorities decided to increase Tests/Case, adding some level of contact tracing? Or conversely, what if they decided to save some costs and make less testing than their current standard of Tests/Case?

Figure 7 shows a simulation using equation (1). For it, I set an arbitrary standard condition at 5 Tests/Cases, attributing to it a death rate of 100%. The graph shows that doubling the number of tests from 5 to 10 makes the death rate fall to 37%, so about one third of the standard condition. Doubling testing rate twice, to 40 Tests/Case, brings death rate to only 5% of the standard condition.

**Figure 7.**
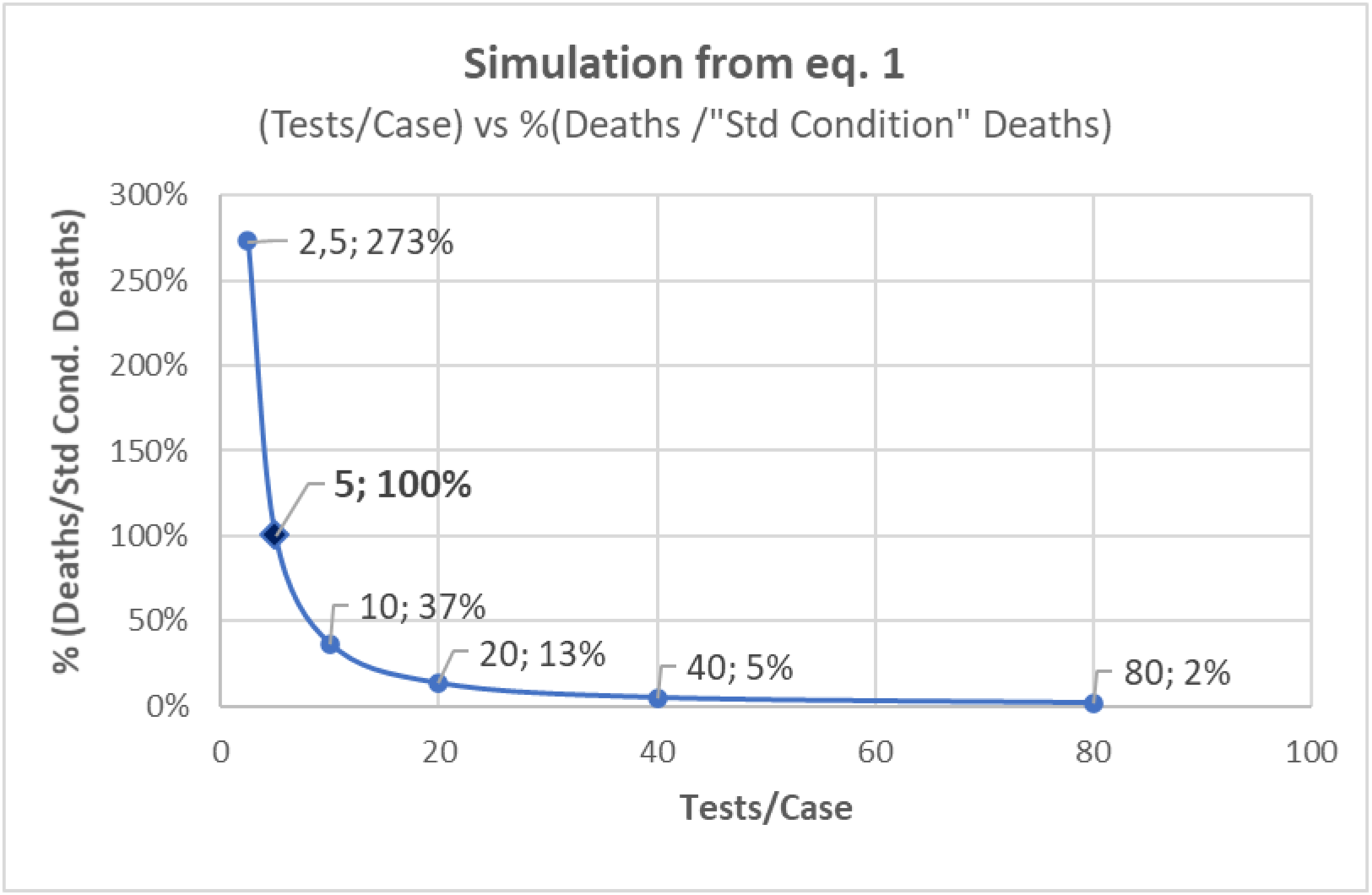
– Simulation from eq. (1) - Effect of Tests/Case on Deaths/Million.

On the other hand, if the health authorities decided to halve their current rate of Tests/Case to 2.5, either for the lack of resources or for they believed they should test only the very sick, the number of deaths would almost triple, reaching 273% of the standard condition.

Those simulations consider just one factor, so they certainly do not capture all the possibilities in pandemic policies. But they show the importance of testing in the context of contact tracing. They help to understand the death rates close to 800 Deaths/Million in New York and New Jersey, both states with less than 3 Tests/Case in the 6^th^ week. For comparison, the state of Washington (13 Tests/Case), Canada (19 tests/Case), Bulgaria (29 Tests/Case) and South Korea (44 Tests/Case) had only 69, 72, 12 and 3,5 Deaths/Million people, respectively.

For the southern countries, the response to increasing testing rate per case was lower, although still remarkably strong. Reasons for this difference between northern and southern countries are unclear and demand further research, but a few possibilities are discussed below.

It is clear by now that pandemics had a slower initial evolution in the southern countries. Even taking Deaths/Million at eight weeks after five deaths, rates are considerably lower than in the northern countries with comparable Tests/Case rates at six weeks.

If the transmission were already more difficult in the southern countries, possible actions to reducing it would become comparatively less important. That would explain the lower impact of Tests/Case variable in the death rate for those countries.

Different percentages of old-age people in the population certainly influenced the pace of the epidemic in those countries. The southern countries studied here have considerably lower percentages of people aged above 65 years, as compared to the northern ones. For instance, the percentages in Peru, Brazil and Argentina are 7,2%, 8,6% and 11,2%, respectively, while being 15,4%, 18,5% and 23% for the USA, UK, and Italy, respectively.

Another obvious hypothesis for a slower transmission rate is the warmer climate in the southern countries. Most of those countries are in tropical and subtropical regions and were hit by the pandemics in late summer or early fall. This hypothesis is supported by a recent study indicating that a warmer climate would help to contain Covid-19 transmission (8), but there is still a lot of controversy about seasonal effects on coronavirus pandemic (9).

We should consider also that we could merely be counting the number of weeks differently. We know by now that Covid-19 was already killing people in Europe by December 2019 (10). In this case the 6^th^ week in Europe could indeed be the 10^th^ week or more. Possibly being hit later made southern countries more aware of the pandemic and allowed earlier detection of cases. Therefore, they would have implemented containment and lockdown earlier and also started counting earlier the pandemic’s weeks.

Regression analysis between Tests/1,000 vs. Deaths/Million showed either a poor or a positive correlation, depending on the set of countries under review. This clearly means that tests per number of inhabitants is a consequence of policies and practices rather than a cause for death rate levels.

A positive correlation between Tests/1,000 and Deaths/Million might indicate an epidemic running out of control in some countries. As the number of cases grows explosively, testing rates, just for the diagnosis of the extremely sick, escalate to quite high Tests/1,000 levels.

In European countries hit early by the pandemic, DIT was the only reasonable choice, as the infrastructure and testing resources were not available to implement a 3T strategy. Among the most populous countries, Germany was the most successful in increasing testing rate and implementing contact tracing. They were making 15 Tests/Case by the 6^th^ week, considerably higher than Italy, Spain and the UK, performing from 4 to 6 Tests/Case. That would explain why Germany had 67 Deaths/Million while similar countries had between 260 to 400 Deaths/Million by the 6th week.

But even Germany’s numbers are dwarfed by the 3,5 Deaths/Million reached by South Korea, that was at the time performing 45 Tests/Case. Or Thailand, which went over 100 Tests/Case and had only 0,8 Deaths/Million by the 6^th^ week after five deaths.

Remarkably, smaller and less wealthy European countries managed to reach high levels of Tests/Case, like Poland, Hungary, Greece, Bulgaria and the Czech Republic. At rates between 25 to 35 Tests/Case, they managed to remain below 35 Deaths/Million.

Sweden is a compelling case in Europe, since they imposed fewer norms of social distancing and kept a higher level of economic activity (11). Besides this, Sweden performed much lower testing than its Scandinavian neighbors. By the 6^th^ week, Sweden was at 6.4 Tests/Case while Denmark, Finland and Norway were at about 20 Tests/Case. As a result, Sweden’s per capita death rate was 3 to 6 times higher than in the other Nordic countries.

Swedish government claimed that they were not seeking for herd immunity without a vaccine. But if they were, their choices would have been quite adequate: limited social distancing, no contact tracing and a low rate of Tests/Case.

Swedish authorities argue that the country’s higher death rate will appear lower in the hindsight (11). The reasoning is that countries following more strict restrictions would suffer further deadly waves of the pandemic when reopening economic activities, while the Swedes would have achieved a certain level of herd immunity.

Claims above deserve to be checked with some facts and mathematics. Swedish authorities made comparisons with European countries that followed DIT strategy. But most European countries are reopening the economy with guidelines for Test & Tracing methodologies, as recommended by the European Union (12) and the OCDE (13). They should though compare the future with, for instance, South Korea’s experience, not with the previous experience in Europe.

Let’s look at some facts and data:

- Korea flattened the epidemic curve in only 20 days after the epidemic outburst, applying social distancing and 3T strategy, but without fully closing business (14). By May 25^th^, three months after the epidemic outburst, they had accumulated only 269 deaths. They had also managed to keep daily deaths below two per day for over one month (average of 0.9 deaths/day). If they succeed in maintain that rate and need to wait one year for a vaccine, they will accumulate 593 total deaths, or 12 Deaths/Million people, by May 2021.
- By the end of May 2020, Sweden already had 400 Deaths/Million people, 34 times the death rate forecasted for Korea one year ahead. And the daily death rate was still in the average of 60 per day, so deaths were still growing significantly. So far, there is no evidence that herd immunity is close in Sweden.

It does not seem necessary to wait to look in hindsight to compare Sweden’s and 3T strategies. Current information already strongly suggests that Sweden’s policy is much less effective in preserving lives, as they have today many more deaths than it is expected for South Korea 12 months ahead.

### 4.2 Some Economic aspects of 3T and DIT strategies

Countries that followed the 3T strategy were able to reopen their economies earlier, while preserving better the health of their citizens. But a detailed economic analysis of different approaches and their impacts on countries’ GDP is outside the scope of this work. I will present here just a brief look at the expenses directly related to the pandemic’s health consequences.

Table 4 shows a simple estimation of the expenses with testing and hospitalization. For the calculations, I assumed the following:

- The unitary cost of PCR tests is US$ 50. That is in line with WHO’s information, placing PCR costs between US$ 30 and US$ 60 per test (15).
- The number of hospitalizations was estimated considering a death rate of 18% for hospitalized people (16). Therefore, the hospitalization rate per million people was taken as 5.5 times Deaths/Million in each country.
- Hospitalizations’ average cost by coronavirus was considered as US$ 4,000 per person, as estimated in a recent paper (17).

**Table 4.** Main direct health expenses caused by the pandemic

| Country | Tests/<br>Case | Testes/<br>1,000 | Deaths/<br>million | Testing Costs/<br>Million (kUS\$) | Hospitalization<br>Costs/ Million<br>(kUS\$) | Total Costs/<br>Million (kUS\$) |
| --- | --- | --- | --- | --- | --- | --- |
| Peru | 1.1 | 2.9 | 88.4 | 147 | 1,945 | 2,092 |
| Ecuador | 1.5 | 2.5 | 160.9 | 126 | 3,540 | 3,666 |
| São Paulo | 3.3 | 13.7 | 102.2 | 685 | 2,248 | 2,933 |
| Chile | 7.7 | 23.1 | 26.6 | 1,156 | 585 | 1,742 |
| Argentina | 11.8 | 2.3 | 8.7 | 117 | 191 | 309 |
| Colombia | 12.6 | 4.4 | 12.1 | 218 | 266 | 485 |
| South Africa | 28.4 | 9.5 | 5.3 | 476 | 117 | 592 |
| Tunisia | 43.4 | 3.8 | 3.98 | 192 | 88 | 279 |
| Thailand | 108.7 | 4.7 | 0.8 | 235 | 18 | 253 |

Table 4 is in crescent order of Tests/Case. As we see, Total Cost/Million people tend to go in the reverse order, meaning that the higher the number of Tests/Case, the lower the total costs.

For instance, São Paulo, with only 3,3 Tests/Case, not only had 20 times more deaths than the lower income South Africa (28 Tests/Case) but was also expending five times more, in per capita basis. São Paulo was still having 12 times more deaths than the very comparable Argentina (12 tests/Case) while expending about ten times more.

Another interesting example is the pair of Tunisia and Ecuador. With close per capita GDP, these middle-low income countries followed quite different strategies. Tunisia was making over 40 Tests/Case by the 6^th^ week and having only 4 Deaths/Million. Ecuador was making only 1.5 Tests/Case but expending 13 times more and having 40 times more deaths per million people.

Figure 8 presents the regression between Tests/Case vs. Costs/Million people for southern countries. Correlation is negative and quite strong, with a determination coefficient equal to 80%. These results dismiss the arguments that 3T is an expensive strategy just affordable by wealthy countries. Not applying it dramatically increases costs and life losses.

**Figure 8.**
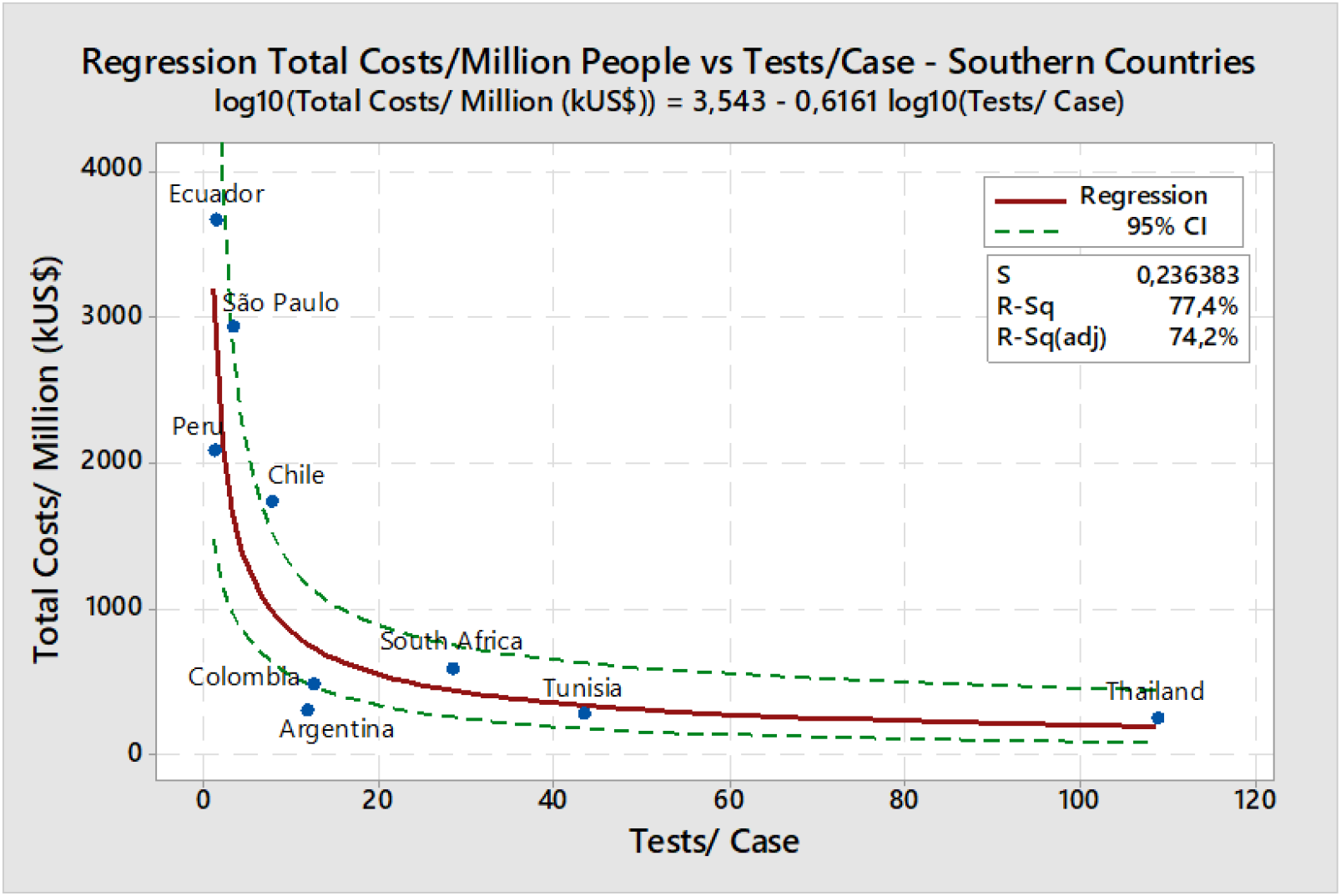
Tests/Case vs Total Costs/Million people – Southern Countries.

A regression between Tests/Case and Deaths/Million was also made for the USA states and Canada. It is clear from the graph in figure 9 that New York and New Jersey, with the lowest levels of Tests/Case, are also the ones with the highest per capita expenses.

**Figure 9.**
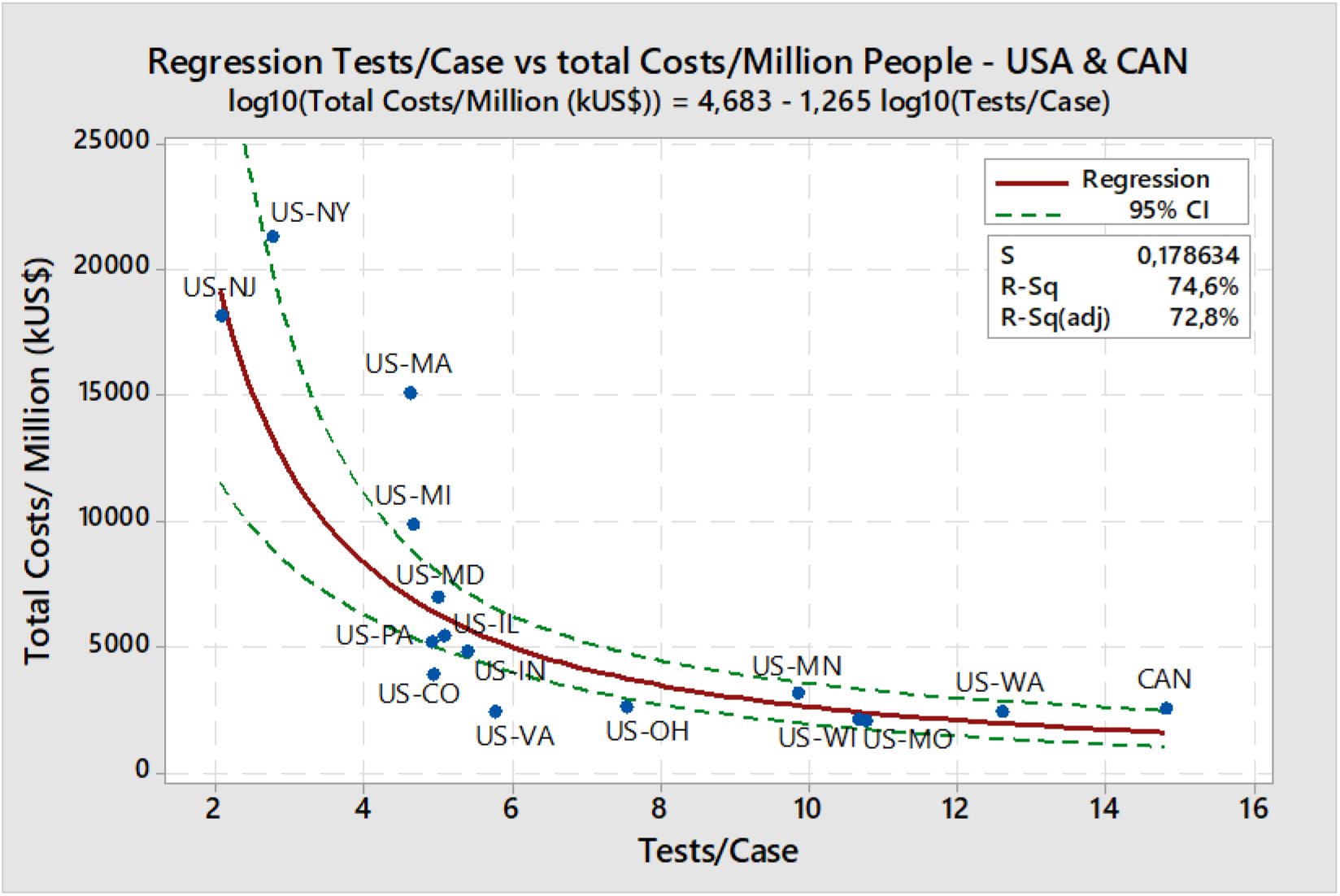
– Tests/Case vs Total Costs/Million people – USA states and Canada.

On the other end, the state of Washington and Canada, with the highest testing per case in the group, not only had the lowest per capita expenses, but also less than half the per capita testing costs found for New York (see table 5).

**Table 5.** Main direct health expenses caused by the pandemic - USA & Canada

| Country | Tests/<br>Case at 6<br>weeks | Testes<br>per 1,000 | Deaths/<br>million | Testing Costs/<br>Million People<br>(kUS\$) | Hospitalization<br>Costs/ Million<br>People (kUS\$) | Total Costs/<br>Million People<br>(kUS\$) |
| --- | --- | --- | --- | --- | --- | --- |
| New Jersey | 2.1 | 27 | 762.4 | 1,359 | 16,773 | 18,131 |
| New York | 2.8 | 41 | 872.3 | 2,049 | 19,190 | 21,239 |
| Massachussets | 4.6 | 46 | 581.1 | 2,283 | 12,785 | 15,068 |
| Michigan | 4.7 | 20 | 402.4 | 1,010 | 8,853 | 9,862 |
| Pennsylvania | 4.9 | 19 | 192.0 | 959 | 4,225 | 5,184 |
| Colorado | 4.9 | 14 | 144.4 | 693 | 3,178 | 3,871 |
| Maryland | 5.0 | 25 | 257.9 | 1,258 | 5,673 | 6,930 |
| Illinois | 5.1 | 22 | 193.9 | 1,123 | 4,266 | 5,389 |
| Indiana | 5.4 | 15 | 182.6 | 774 | 4,018 | 4,791 |
| Virginia | 5.8 | 13 | 80.1 | 657 | 1,762 | 2,419 |
| Ohio | 7.5 | 13 | 88.8 | 642 | 1,953 | 2,596 |
| Minnesota | 9.8 | 19 | 98.9 | 942 | 2,177 | 3,118 |
| Missouri | 10.7 | 15 | 61.4 | 774 | 1,351 | 2,125 |
| Wisconsin | 10.8 | 15 | 58.4 | 762 | 1,285 | 2,047 |
| Washington | 12.6 | 18 | 69.1 | 876 | 1,521 | 2,397 |
| Canada | 14.8 | 19 | 71.8 | 952 | 1,580 | 2,532 |

It is also apparent from Table 5 that the major costs are the hospitalization ones. For New York, hospitalization costed about 10 times more than testing. For Washington, hospitalization was less than twice the costs for testing, which in turn was about 13 times lower than for New York, in per capita basis. That is of course a consequence of the lower number of cases and deaths.

Regression equation shown in figure 9 indicates that doubling Tests/Case reduces total per capita costs by a factor of 2,4 times, for this group of “countries”. For the set of southern countries, regression indicated a lower impact, but even so, doubling Tests/Case would divide expenses by 1.5 times.

The conclusion is that 3T-like strategies should not be considered as expensive and unaffordable ones, as they indeed reduce overall costs, while protecting people’s health and lives.

### 4.3 A further look at the Americas

Despite being hit later, most of the countries in the Americas are not following 3T strategy or doing high levels of testing per case. This contrasts with countries like South Africa (28 Tests/Case), Tunisia (40 Tests/Case) and Thailand (108 Tests/case) which are doing contact tracing and having below five deaths per million people, despite not being particularly wealthy and resourceful countries.

It seems that a lot of misunderstandings are preventing the recognition of 3T advantages in the Americas. In a recent press briefing (18), the government of the USA compared their testing rates with the ones in Korea, claiming to have twice as much per capita testing, so doing much better than that country.

Despite the high level of per capita testing, the USA had an average of 9,7 Tests/Case by the 6^th^ week but had started with a considerably lower rate. South Korea’s half per capita testing rate was applied in the context of 3T strategy (45 Tests/Case) and brought them to 2% of the US per capita death rate. They just did not need to do more testing, since the epidemic had already been crushed.

Possibly more than any other country, high per capita testing in the USA is a consequence of an epidemic that went out of control, with explosive growth in the number of cases. That is suggested by the strong and positive correlation between Tests/1,000 and Deaths/Million when dealing with the USA northern states and Canada data alone.

In Brazil, the federal government rejected the recommendations of WHO on intensive testing and tracing (19), deciding to test only those seriously sick. The Health Minister declared that it was unfeasible to test 100% of the population, a waste of public resources, mischaracterizing the 3T strategy. The Health Ministry also stated that they intended to build immunity, with about 50% of the population contaminated by September 2020 (20). Those approaches, combined with the Brazilian President’s denial about the pandemics and sabotaging mitigation initiatives by the state governors, are pushing Brazil to a critical situation (21).

Despite the fast growth in cases, Brazilian government decisions on testing were still in place by May 2020 and Brazil was making about three tests per case. So far, there are no studies on implementing contact tracing, neither by the Health Ministry nor by the governments of the states.

In a recent paper by Eichenbaum et al (22) about the effects of testing and quarantining on the pandemic, the authors conclude that high testing would bring the best tradeoff between economic performance and health outcomes. In the paper’s context, increased testing is made to allow for contact tracing, so their modeling seems compatible with the findings in this work.

For countries lacking capacity of testing for tracing, or which just need to reduce tracing costs, the use of the so called “group testing” or “pool testing” has been proposed (23, 24). Laboratory methodologies have already been developed, as described in a few papers (25, 26, 27).

## 5 Conclusions

The affirmation that 3T is a “mass testing” strategy must be analyzed in the right context. It is “mass testing”, as it starts at a high daily testing rate, while DIT begins at a much slower pace. But daily numbers tend to grow exponentially with time for DIT, while for 3T they remain relatively constant or decay, as the daily number of cases fall quite fast. In the bottom line, very often DIT is in fact the “mass testing” strategy, with the highest levels of per capita testing.

Correlation studies here presented indicate that Tests/Case is a good predictor for the death rate and a better parameter to evaluate policies against the pandemic. On the other hand, tests per number of inhabitants is a poor predictor and could even be an indicator of an epidemic that ran out of control, with the exponential growth of cases and deaths.

Claims that 3T is more expensive than DIT and unaffordable by poorer countries are wrong. If applied in an early stage, 3T would indeed reduce the expenses, while sparing many lives.

While in Europe “Test & Tracing” is becoming a mantra for the opening phase of the pandemics, the base for the “new normal”, the concept has not been accepted in most parts of the Americas.

Mitigation strategies are not enough to maintain an acceptable compromise involving health outcomes, people’s quality of life and economic performance in the longer term. A better alternative is to combine (shorter) lockdowns with containment strategies like the 3T one, which should allow acceptable compromises until a Covid-19 vaccine is available to promote herd immunity.

## Data Availability

All data is available in the databases of Worldometer, Our World in Data and The COVID Tracking Project websites.

https://www.worldometers.info/coronavirus/

https://covidtracking.com/

